# Drug-induced liver injury associated with elexacaftor/tezacaftor/ivacaftor from the FDA Adverse Event Reporting System (FAERS)

**DOI:** 10.1101/2023.09.16.23295574

**Authors:** Alan Shi, Harold Nguyen, C. Benson Kuo, Paul M. Beringer

## Abstract

**Introduction:** The efficacy and safety of elexacaftor/tezacaftor/ivacaftor (ETI) have been established in prospective clinical trials. Liver function test elevations were observed in a greater proportion of patients receiving ETI compared with placebo; however, the relatively small number of patients and short duration of study preclude detection of rare but clinically significant associations with drug-induced liver injury (DILI). To address this gap, we assessed the real-world risk of DILI associated with ETI through data mining of the FDA Adverse Event Reporting System (FAERS).

**Methods:** Disproportionality analyses were conducted on FAERS data from the fourth quarter of 2019 through the third quarter of 2022. Comparative patient demographics, onset time and outcomes for ETI-DILI were also obtained.

**Results:** 452 reports of DILI associated with ETI were found, representing 2.1% of all adverse event reports for ETI. All disproportionality measures were significant for ETI-DILI at p < 0.05; the reporting odds ratio (ROR) was comparable to that of drugs classified by FDA as “Most-DILI concern”. The most notable demographic finding was a male majority for ETI-DILI compared to a female majority for non ETI-DILI. Median ETI-DILI onset time was 50.5 days, and hospitalization was the second most common complication.

**Conclusion:** Using FAERS data, ETI was found to be disproportionality associated with DILI. Future research is needed to investigate the hepatotoxic mechanisms and assess potential mitigation strategies for ETI-induced hepatotoxicity.

**Article Highlights:**

- Using the FDA Adverse Event Reporting System database, ETI and DILI were found to be significantly associated (p < 0.05) for all disproportionality measures (PRR, ROR, IC, EGBM, Yates’ chi-squared).
- The ROR for ETI-DILI is greater than that of many “Most-DILI concern” drugs in the FDA DILIRank dataset but is not within the top 20 drugs associated with DILI.
- Patient reports for ETI-DILI were predominately male, in contrast to patient reports for other drugs and DILI.
- “Hospitalization” was the second most common patient outcome for ETI-DILI after “other serious outcomes”.
- Most patients had onset times within 3 months of initiation, several patients had an onset time greater than 1 year.
- Onset times indicate that liver function test monitoring should be initiated earlier than 3 months and potentially extend beyond 1 year in some patients.

## 1. Introduction

Elexacaftor/tezacaftor/ivacaftor (ETI, brand name: Trikafta) is a cystic fibrosis transmembrane conductance regulator (CFTR) modulator which acts as a triple combination protein corrector-potentiator therapy. Clinical trials have shown that ETI is highly efficacious in improving lung function and nutritional status in people with cystic fibrosis and is well tolerated with few safety concerns (1-6). However, periodic monitoring is recommended due to the increased incidence of liver function test abnormalities identified in clinical trials as well as in case reports of drug-induced liver injury (DILI) (1-8). DILI due to ETI can be predicted in part by its physicochemical properties. The Biopharmaceutical Classification System (BCS) classifies drugs according to permeability and solubility. Among the compounds in ETI, ivacaftor and tezacaftor fall under the Biopharmaceutical Classification System (BCS) Class 2, based on high permeability and low solubility (9, 10). Compounds in Class 2 of BCS and the similar Biopharmaceutical Drug Disposition Classification System (BDDCS) are considered to have the highest DILI risk compared to other classes (11).

Consistent with the predicted DILI risk for BCS Class 2, Phase III clinical trials showed evidence of DILI due to ETI, including elevation in ALT/AST > 5X the upper limit of normal (ULN) or ALT/AST > 3X ULN with bilirubin > 2X ULN in the absence of other causes such as hepatitis or alcoholic liver disease (1-6). Results from one of the pivotal phase 3 trials showed that the incidence of maximum transaminase elevations above 8X, 5X, or 3X the upper limit of normal (ULN) occurred in 1%, 2%, and 8% of ETI-treated patients compared to 1%, 1%, and 5% for placebo, while the incidence of maximum total bilirubin above 2X ULN occurred in 4% of ETI-treated patients compared to <1% for placebo (1). In real-world use, DILI occurring after ETI administration has resulted in several cases of severe outcomes for patients, including acute liver failure requiring transplantation and a case of hepatic necrosis (12-14). A limitation of Phase III trials is their relatively small sample size and short term follow up which cannot capture rare safety concerns that may be revealed in a longer-term, larger pool of real-world patients.

Despite the predictive risk of DILI due to ETI’s physicochemical properties and evidence of DILI associated with ETI in clinical trials and post-marketing surveillance, a statistical association between ETI and DILI in real-world use has not been established. While the FDA has created DILIrank, a dataset of 1,036 FDA-approved drugs divided into four classes based on their potential for causing DILI, elexacaftor, tezacaftor, and ivacaftor are noticeably absent from this vital resource (15). The analysis of large adverse event databases, such as the FDA Adverse Event Reporting System (FAERS), can improve understanding of the ETI-DILI relationship by detecting and determining the strength of disproportionality signals for ETI-DILI and identifying DILI-related safety concerns of ETI that may be missed in clinical trials and case reports. This study aims to shed light on the real-world risk of DILI associated with ETI along with relevant demographic and patient outcome trends related to ETI-DILI through the data mining of FAERS quarterly data reports three years from the approval of ETI in the US market, from Q4 2019 to Q3 2022, inclusive (16).

## 2. Methods

### 2.1 Source of Data

The FDA Adverse Event Reporting System (FAERS) database supports FDA’s post-marketing surveillance for drugs and biologics and contains reports of adverse events, medication errors, and product quality complaints (17). Reports in FAERS may be submitted by healthcare professionals, consumers, and manufacturers to the MedWatch website to be compiled in FAERS (17). FAERS is structured into several datasets containing drug information (DRUG), demographic information (DEMO), patient outcomes (OUTC), indications (INDI), adverse reactions (REAC) and drug therapy start dates and end dates (THER). The datasets are linked through a primary identification number (PRIMARYID). Definitions of the outcome codes from the MedWatch form instructions are included in Supplement S1 (18).

FAERS data from the fourth quarter of 2019 through the third quarter of 2022 were analyzed using the data analytics platform, KNIME (Version 4.7.3), and Microsoft Excel (16, 19). Specifically, the DEMO, THER, INDI, DRUG, and REAC files were utilized to formulate a contingency table and OUTC was used to analyze outcome data. The data analysis process involved file merging, deduplication, categorization of the reports into a contingency table, and post-grouping statistical analysis. A PDF of the KNIME data analysis workflow can be found in Supplement S2, and the KNIME workflow files can be found in Supplement S3.

### 2.2 File Merging

The 12 quarters of data for each of the DEMO, THER, INDI, DRUG, REAC, and OUTC .txt files were merged using the command prompt in Windows. The merged DEMO, THER, INDI, DRUG, REAC, and OUTC .txt files were uploaded to KNIME and the rows with column headers such as “primaryid” were removed with a row filter exclusion. The files were then converted to CSV using KNIME to begin the deduplication process. Duplicates may occur because of redundant reports submitted by different sources (such as drug manufacturers, patients, and healthcare providers), and reoccurrences of the same report across multiple quarters due to follow-ups of the original case.

### 2.3 Deduplication

Cases in the FAERS database are represented by a numerical CASEID and a numerical PRIMARYID, a combination of the CASEID and Case Version number. The first deduplication step involves keeping only the latest PRIMARYID for any set of CASEID duplicates. The second deduplication step (based on Khaleel et al., 2022) is performed by removing event entries that match in all the following 8 categories: event date, patient age, patient sex, reporter country, list of start dates, list of indications, list of active ingredients, and list of preferred terms of adverse events (20). In other words, two reports will be considered as one if they match in the above 8 categories, even if they possess distinct CASEIDs. Note that reports lacking age data are treated as distinct entities.

As the DEMO data has unique PRIMARYIDs, deduplication began with this database, then the deduplicated DEMO was combined with the other data sets to eliminate all duplicate PRIMARYIDs. Sorted lists of start dates, indications, active ingredients, and preferred terms are created with GroupBy nodes and merged with the DEMO data to allow for Duplicate Row Filter to find additional duplicates. Finally, any duplicates from this filter that are missing age are added back into the pool of events.

The deduplicated unique data was used to create a contingency table that categorizes each adverse reaction into one of four categories: ETI-DILI, ETI-not DILI, not ETI-DILI, not ETI-not DILI. To search for entries that include our drug of interest, several nodes were used to find active ingredients (the prod_ai column in the FAERS data) that matched “ELEXACAFTOR/IVACAFTOR/TEZACAFTOR” or “ELEXACAFTOR”, or drug names matching “TRIKAFTA” or “KAFTRIO”.

Prior to the drug search, the REAC dataset was modified by replacing related liver injury preferred terms to “DILI”. Table 1 lists a dictionary of preferred terms for liver injury from the Medical Dictionary for Regulatory Activities (MedDRA), obtained from a prior study investigating antifungal drugs and DILI (21). Multiple instances of “DILI” per patient, resulting from the patient having multiple preferred terms that translate to “DILI”, were removed from the sample. This list of preferred terms was used for the deduplication of DEMO. The list of translated reactions is cross referenced with the list of ETI matches, leading to a completed contingency table, shown in Table 2.

**Table 1.** Dictionary of Preferred Terms for Liver Injury (21)

| Preferred Terms (PT) |
| --- |
| Alanine aminotransferase abnormal |
| Alanine aminotransferase increased |
| Ammonia increased |
| Aspartate aminotransferase abnormal |
| Aspartate aminotransferase increased |
| Bile duct damage |
| Biliary cholangitis |
| Bilirubin conjugated increased |
| Bilirubin urine |
| Blood bilirubin abnormal |
| Blood bilirubin increased |
| Blood bilirubin unconjugated increased |
| Cholestasis |
| Cirrhosis |
| Coma hepatic |
| DILI |
| Drug-induced liver injury |
| Hepatic cirrhosis |
| Hepatic damage |
| Hepatic disease |
| Hepatic encephalopathy |
| Hepatic enzyme abnormal |
| Hepatic enzyme increased |
| Hepatic failure |
| Hepatic function abnormal |
| Hepatic injury |
| Hepatic necrosis |
| Hepatic steatosis |
| Hepatic vascular injury |
| Hepatitis |
| Hepatobiliary disease |
| Hepatocellular damage |
| Hepatocellular injury |
| Hepatomegaly |
| Hepatopathy |
| Hepatotoxicity |
| Hyperammonaemia |
| Hyperbilirubinaemia |
| Icterus |
| Jaundice |
| Liver damage |
| Liver fatty infiltration |
| Liver function test abnormal |
| Liver injury |
| Liver necrosis |
| Liver transplant |
| Mixed hepatocellular-cholestatic injury |
| Nonalcoholic fatty liver disease |
| Portal hypertension |
| Steatohepatitis |
| Transaminases abnormal |
| Transaminases increased |
| Urine bilirubin increased |

**Table 2.** Contingency table and disproportionality analysis for ETI-induced DILI.

|  | **ETI** | **Not ETI** | **Total** |
| --- | --- | --- | --- |
| **DILI** | 452 (A) | 86268 (B) | 86720 (A + B) |
| **Not DILI** | 20537 (C) | 13469463 (D) | 13490000 (C + D) |
| **Total** | 20989 (A + C) | 13555731 (B + D) | 13576720 (A+ B + C + D) |

| Measure of Disproportionality | Value |
| --- | --- |
| PRR (95% CI) | 3.38 (3.08, 3.71) |
| ROR (95% CI) | 3.44 (3.13, 3.77) |
| EBGM (95% CI lower bound) | 3.37 (3.12) |
| Yates' Chi-squared | 757.61 |
| IC (95% CI) | 1.75 (1.59, 1.86) |

### 2.4 Demographic Extrapolation

Once categorized into ETI-DILI, ETI-not DILI, not ETI-DILI, not ETI-not DILI, the adverse reaction reports were grouped for each patient to collect demographic data, as one PRIMARYID can correspond to multiple adverse events, start dates, end dates, and outcomes. Thus, a patient could contribute to the statistics of multiple contingency table groups. Ages and weights were converted to all have units of years and kilograms, respectively, based on their corresponding codes. When calculating descriptive statistics for age and weight, cutoffs of 0-115 years of age and 0-300 kg weight were introduced to exclude biologically implausible values that may have been entered erroneously. Outcome codes were also tabulated for the patients, counted as unique instances of each code (a patient could have both “Other Serious Outcome” and “Hospitalization” counted, but two “Hospitalization” events would be counted once). Definitions of the outcome codes from the instructions for completing MedWatch forms are included in Supplement S1 (18). Onset times for events were calculated only for Group A and only if a full date (YYYYMMdd) was available for event date and start date, with a cutoff minimum date of 20190101.

### 2.5 Statistical Analysis

Several quantitative measures of correlation between ETI use and DILI reports were used, including proportional reporting ratio (PRR), reporting odds ratio (ROR), Yates’ chi-squared test (χ^2^_yates_), the information component (IC), and the empirical Bayesian geometric mean (EBGM).

PRR is a measure of the disproportionality of reporting of an adverse event for a product of interest in comparison to the same adverse event for all other products in the database. ROR is the odds of an adverse event occurring for a product of interest compared to the odds of that event occurring for all other products in the database. EGBM is a Bayesian statistical analysis that is similar to PRR but produces disproportionality scores toward the null. For PRR, ROR, and EBGM, statistically significant signals occur when the lower limit of the 95% confidence interval (CI) is greater than 1. Yates’ chi-squared test measures the degree of difference between the observed and expected number of reports for a drug-event combination, corrected for the assumption that the binomial frequencies from the contingency table can be approximated by the continuous chi-square distribution. The minimum criteria for adverse event safety signals established by Evans et al. (2002) is a PRR of at least 2, chi-squared of at least 4, and 3 or more or cases (22).

IC measures the disproportionality between the observed and expected number of reports for a drug-event combination, such as ETI-DILI for this study. Positive IC results from a higher number of observed reports than expected reports, while negative IC results from a lower number of observed reports than expected reports. Statistically significant signals occur when the 95% CI does not contain 0.

## 3. Results

### 3.1. Descriptive Analysis

There were 13,576,720 adverse reaction reports considered after deduplication, yielding 20,989 reports containing ETI in the associated drug list. Among the ETI reports, 452 (2.1%) were considered DILI related. A contingency table of the reports and Bayesian analysis metrics are shown in Table 2. Demographic data on the categorized reports are shown in Table 3. Most patients in the intersection of ETI and DILI were male (61% vs 37%), while the other adverse reactions had more females than males.

**Table 3.** Demographic Data of Categorized Events.

|  | ETI, DILI<br>(A) | ETI, Not<br>DILI (C) | Not ETI, DILI<br>(B) | Not ETI, Not<br>DILI (D) |
| --- | --- | --- | --- | --- |
| **N (patients)** | 452 | 8569 | 86268 | 4522904 |
| **Sex (n)** |  |  |  |  |
| F | 168 (37%) | 4601 (54%) | 42703 (50%) | 2291612 (51%) |
| M | 240 (53%) | 3421 (40%) | 32392 (38%) | 1632874 (36%) |
| Blank | 44 (10%) | 546 (6%) | 11164 (13%) | 597008 (13%) |
| Other (U, P, I, T) | 0 (0%) | 1 (0%) | 9 (0%) | 1410 (0%) |
| **Age (years)** |  |  |  |  |
| Median (IQR) | 27 (16-38) | 26 (18-37) | 57 (42-68) | 59(43-70) |
| Mean [SD] | 28.86<br>[14.99] | 29.04 [14.71] | 53.31 [19.93] | 55.32 [20.11] |
| Missing or Excluded | 187 | 3933 | 26510 | 2088609 |
| **Weight (kg)** |  |  |  |  |
| Median (IQR) | 58 (49-68) | 59 (50-70) | 70 (57-85.5) | 73 (60-89) |
| Mean [SD] | 58.86<br>[15.72] | 60.83 [17.82] | 71.58 [25.11] | 75.19 [26.19] |
| Missing or Excluded | 307 | 6160 | 60075 | 3762110 |
| **Age Distribution (n)** |  |  |  |  |
| <0 (excluded) | 0 (0.00%) | 0 (0.00%) | 0 (0.00%) | 21 (0.00%) |
| 0-18 | 75<br>(16.59%) | 1083<br>(12.64%) | 4057 (4.70%) | 141207 (3.12%) |
| 18-29 | 70<br>(15.49%) | 1607<br>(18.75%) | 3855 (4.47%) | 170252 (3.76%) |
| 30-49 | 97<br>(21.46%) | 1434<br>(16.73%) | 14165 (16.42%) | 490508 (10.84%) |
| 50-64 | 14 (3.10%) | 378 (4.41%) | 18358 (21.28%) | 723954 (16.01%) |
| 65-74 | 7 (1.55%) | 103 (1.20%) | 11903 (13.80%) | 510356 (11.28%) |
| 75-84 | 2 (0.44%) | 30 (0.35%) | 5859 (6.79%) | 305606 (6.76%) |
| 85-115 | 0 (0.00%) | 1 (0.01%) | 1561 (1.81%) | 92412 (2.04%) |
| >115 (excluded) | 0 (0.00%) | 0 (0.00%) | 1 (0.00%) | 46 (0.00%) |
| Missing | 187<br>(41.37%) | 3933<br>(45.90%) | 26509 (30.73%) | 2088542 (46.18%) |

### 3.2 Disproportionality Analysis

The PRR was 3.38 (95% CI: 3.08-3.70), indicating an increased reporting rate of DILI with ETI use compared to DILI with non-ETI drugs. The ROR was 3.44 (95% CI: 3.13-3.77), also suggesting an increased likelihood of ETI patients experiencing DILI compared to all other drugs. Similarly, the EBGM of 3.37 (95% CI lower bound: 3.12) also showed a signal for increased reporting of DILI while taking ETI.

## 4. Conclusion

Based on the data from the FDA Adverse Event Reporting System database, ETI has been shown to be significantly associated with DILI for all the given measures of statistical disproportionality, including PRR, ROR, IC, EGBM, and Yates’ chi-squared. The lower bound of the 95% confidence interval for the PRR, ROR, and IC of the ETI-DILI association were all greater than 1 (and greater than 0 in the case of IC), indicating that the association between ETI and DILI is statistically significant with p < 0.05. The Yates’ chi-squared statistic of 757.61 was far greater than the critical value (10.828) for 1 degree of freedom at p < 0.001, indicating a highly significant association between ETI and DILI. Most importantly, ETI and DILI clearly exceeded the minimum requirement for adverse event safety signals (PRR ≥ 2, chi-squared ≥ 4, n ≥ 3) (22).

### 4.1 Demographics

The demographic data of the contingency table reveal notable differences between the four groups. The proportion of male to female patients for the ETI-DILI group (53% M, 37% F) is higher than that the ETI-not DILI group (40% M, 54% F), the not ETI-DILI group (38% M, 50% F), and the not ETI-not DILI group (36% M, 51% F). The higher proportion of males to females (3:2) in group the ETI-DILI group is notable because it runs counter to the evidence that women have been found to be more susceptible than men to DILI (23). Comparing the age across groups revealed that the ETI groups tended to be younger (median age 27 and 26, respectively) than non-ETI groups (median age 57 and 59, respectively), consistent with cystic fibrosis being detected in children and being treated as early as possible. Patient weight followed a similar pattern: the ETI groups tended to have lower weight (median weight 58 kg and 59 kg, respectively) than non-ETI groups (median weight 70 kg and 73 kg, respectively). The lower weight in ETI groups may be due to the younger age of the ETI groups and/or cystic fibrosis-related malnutrition that has not been fully offset by ETI.

### 4.2 Monitoring

The calculated onset time of DILI for ETI and the distribution of outcomes from the two ETI groups (A and C) shows that DILI tended to occur in patients less than 6 months after taking ETI and that a significant portion of patients required hospitalization. The calculated median onset time of 50.5 days (interquartile range: 18 - 149 days) and the mean of 113.3 days (SD = 155.7 days) suggest that the typical onset time for DILI after ETI administration is approximately 2 to 4 months, matching the package insert guidance to monitor for elevated liver function tests every 3 months during the first year of treatment (24). However, most patients (62%, 72 out of 116) had an onset time less than 3 months (90 days) and many patients (41%, 48 out of 116) had an even shorter onset time less than one month (30 days). A notable minority of patients (7%, 8 out of 116) experienced onset times greater than 1 year, with the longest being over 800 days (Figure 2). Thus, the data for early-onset and late-onset DILI due to ETI suggests that clinicians should monitor for DILI within one month after starting ETI and should remain vigilant for rare but potentially significant DILI events past the one-year window, since a significant portion of these events resulted in hospitalization. “Hospitalization” was the second most common patient outcome at a rate of 38% among the ETI-DILI group and 32% among the ETI-non DILI group (Table 4). Compiling the top 20 adverse reactions of hospitalized patients in the ETI-DILI group revealed that the top 4 adverse reactions were preferred terms for DILI, while the 5^th^ was “infective pulmonary exacerbation of cystic fibrosis” (Table 5). Other non-DILI adverse reactions were also part of the top-20 list, suggesting that the high rate of hospitalization was not exclusively due to ETI-induced DILI.

**Table 4.** Complications for Patients Receiving ETI.

| Outcome | ETI, DILI | % patients with outcome | ETI, Not DILI | % patients with outcome |
| --- | --- | --- | --- | --- |
| Missing | 205 | 45% | 4675 | 55% |
| Congenital Anomaly | 2 | 1% | 26 | 0% |
| Death | 13 | 4% | 171 | 2% |
| Disability | 1 | 0% | 30 | 0% |
| Hospitalization | 124 | 38% | 2727 | 32% |
| Life-threatening | 7 | 2% | 38 | 0% |
| Other Serious Outcome | 193 | 60% | 1604 | 19% |
| Required Intervention | 0 | 0% | 4 | 0% |
| Total | 545 | N/A | 9275 | N/A |

**Table 5.**
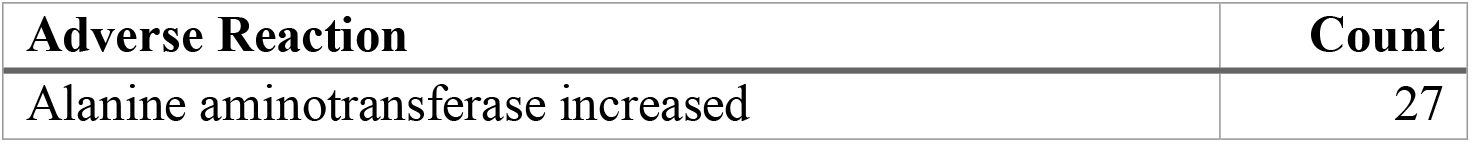
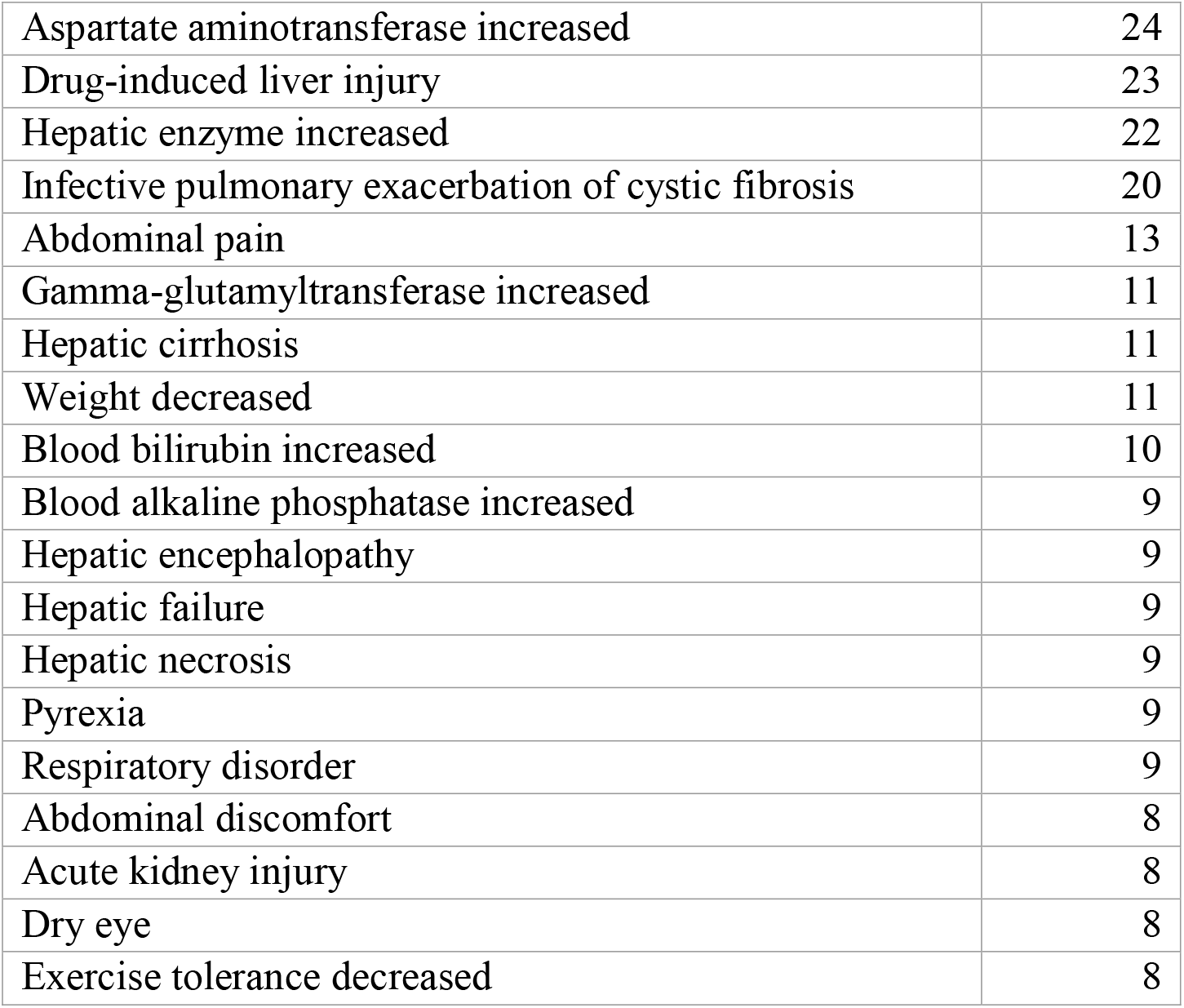
Top 20 Adverse Reactions of Hospitalized Patients in Group A (ETI + DILI)

**Figure 1.**
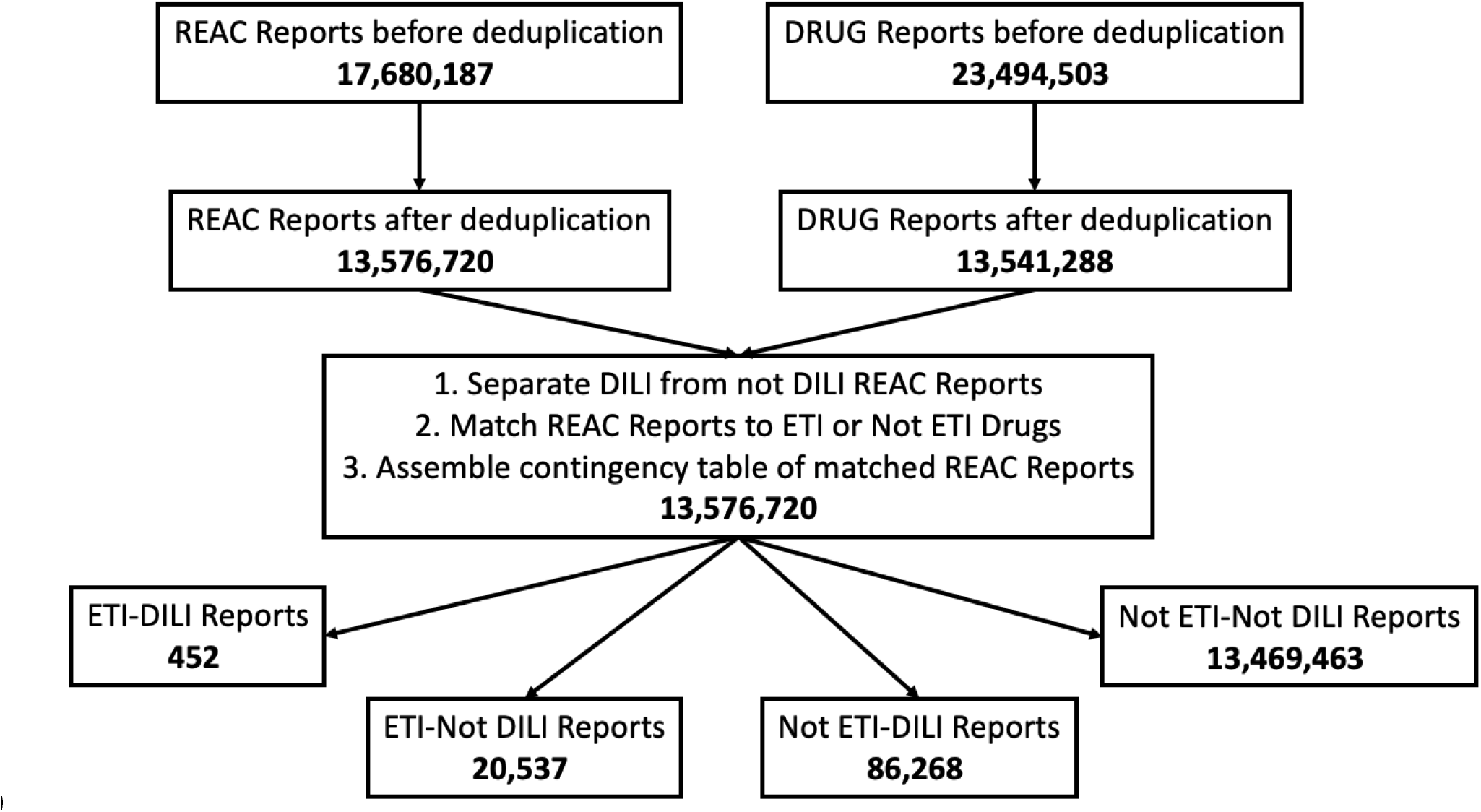
Flowchart of FAERS data manipulation steps with the number of reports shown in bold.

**Figure 2.**
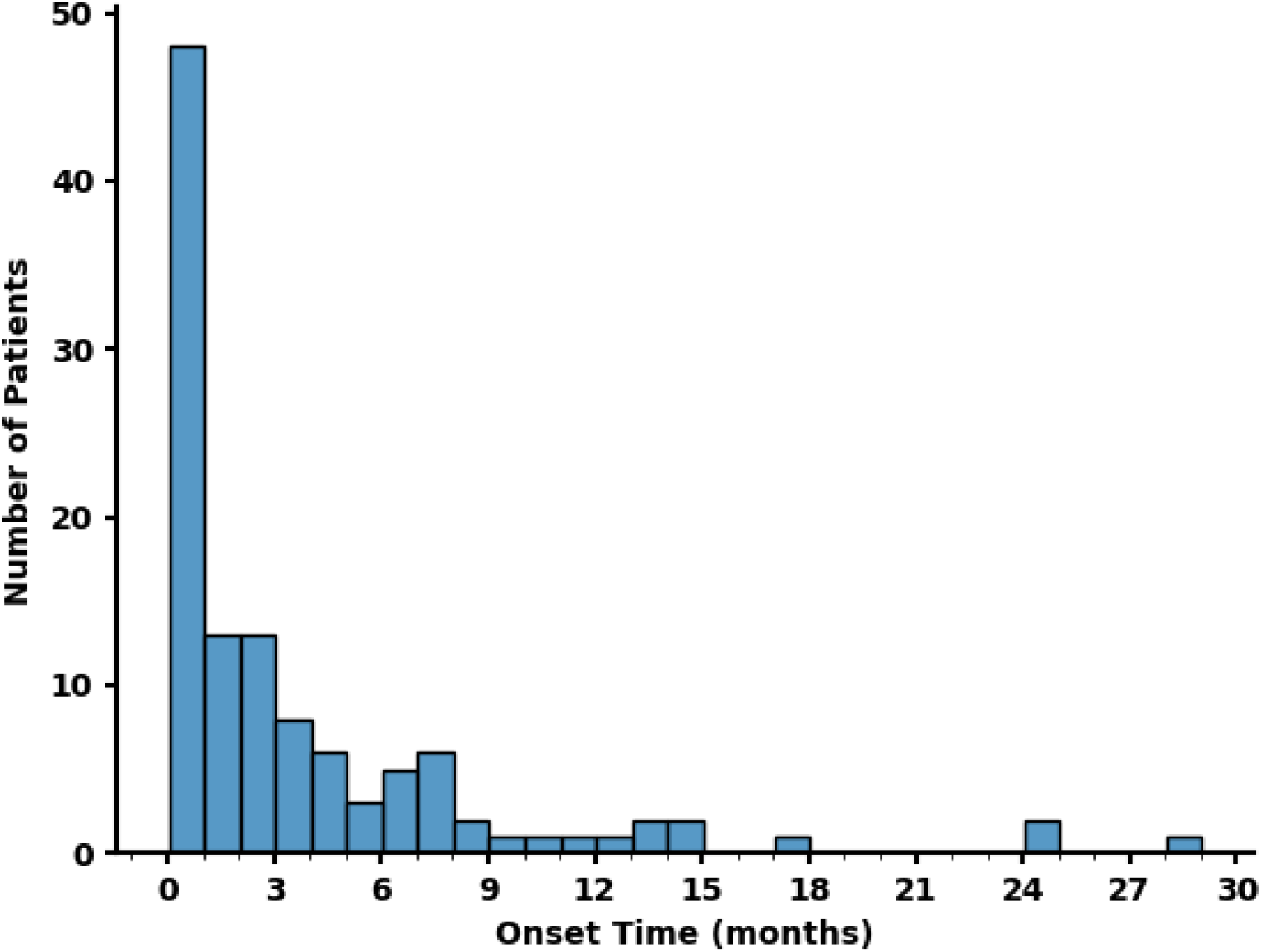
Frequency distribution of onset time for DILI after ETI initiation.

### 4.3 Comparison with Other Drugs

The ROR for ETI and DILI is greater than that of many drugs among the 192 classified as “Most-DILI concern” in the National Center for Toxicological Research Liver Toxicity Knowledge Base (NCTR-LTKB) DILIrank dataset (15, 25). This aligns with the prediction of DILI risk from BDDCS, as Class 2 drugs such as ETI represent 53.6% of the “Severe DILI” category (acute liver failure, fatal hepatotoxicity, and discontinued and withdrawn), while Class 1, 3, and 4 represent 28.6%, 14.3%, and 3.6%, respectively (11). In addition, the high dosage and extensive hepatic metabolism (≥ 50%) of ETI are factors identified by FDA as strongly associated (p < 0.05) with increased DILI risk (26). Nonetheless, the ROR of ETI-DILI, 3.44 (95% CI: 3.13 – 3.77), is still lower than that of many drugs currently on the market that are prescribed for people with cystic fibrosis, such as rifampin, 18.64 (17.54 – 19.82), atorvastatin, 4.479 (4.358 – 4.604), clarithromycin, 4.809 (4.538 – 5.096), itraconazole, 5.552 (5.097 – 6.048), voriconazole, 6.229 (5.79 – 6.702), and ciprofloxacin, 3.272 (3.095–3.459). In all, although ETI’s ROR of 3.44 surpasses that of numerous drugs in the “Most-DILI concern” category in DILIrank, it does not rank in the top 20 drugs associated with DILI (25).

### 4.4 Limitations

Limitations of the study include incomplete data from the FAERS database. For example, across all 4 groups of the contingency table, 6-13% of the patients’ sex was left blank. In addition, it was common for patient reports in the database to lack a recorded outcome measure, as evidenced by missing outcomes for 43% of patients in the ETI-DILI group and 55% of patients in the ETI-not DILI group. There may also be other potential covariates that could cause DILI in the ETI-DILI group, including coadministered drugs (such as acetaminophen) and preexisting liver disease/other conditions, as well as factors not available in FAERS, such as alcohol/illicit drug use. Therefore, data from FAERS can only be used to assess the association between ETI and DILI, not causation (27). Despite these limitations, the data show that ETI and DILI are disproportionally associated compared to all other drugs and all other adverse reactions; the risk of DILI for ETI with or without other drugs is significantly higher than the overall risk of DILI for all other drugs in FAERS.

### 4.5 Novelty of KNIME

Traditionally, script-based programming languages such as Python and SAS have been used to screen and analyze the FAERS database. For example, Python has been used to analyze FAERS data for a pharmacovigilance study of SARS-CoV-2 therapies and SAS has been used to mine FAERS data to determine the statistical association between dipeptidyl peptidase-4 and venous thromboembolism (28, 29). Though they are effective and efficient tools to conduct data mining studies, the need to be proficient in programming results in a high entry barrier for analyzing databases such as FAERS. Alternatively, visual programming software would be more preferrable and appropriate for a broader scientific audience. One such program is KNIME, the Konstanz Information Miner, a free and open-source data analysis platform. KNIME has been used to analyze databases other than FAERS for a variety of applications within pharmaceutical and life sciences, such as searching the Protein Data Bank to support ligand- and structure-based drug design, and parsing genomic reference databases to identify and quantify microorganisms from the metagenomes of microbial communities (30, 31). To our knowledge, this study has been the first use of KNIME for performing disproportionality analyses on adverse event data from the FAERS database. Our KNIME workflow is available in Supplement S3 and can be adapted and reused to find association of other drugs with DILI.

### 4.6 Summary and Future Directions

In summary, data mining the FAERS database using KNIME found that the association between ETI and DILI is statistically significant across all the major measures of disproportionality analysis, including PRR, ROR, IC, EBGM, and Yates’ chi-squared. The ROR of the ETI-DILI association would place ETI among the 192 drugs listed as “Most-DILI concern” in the NCTR-LTKB DILIrank dataset, but not within the top 20 drugs on the list. Nevertheless, the high rate of hospitalization among the ETI-DILI group in the FAERS data underscores the necessity to carefully monitor cystic fibrosis patients taking ETI and initiate interventions to avoid DILI. While the association between ETI and DILI has been established from real-world data in FAERS, future studies are needed to establish the causal relationship between ETI and DILI by investigating the mechanisms of ETI-induced DILI. Additional research is also needed to assess potential mitigation strategies including ETI dose reduction and/or discontinuation to determine the efficacy and safety of these approaches (32).

## Data Availability

All data produced in the present study are available upon reasonable request to the authors.

## Author Contributions

Conceptualization, A.S. and P.M.B.; Data curation, A.S. and H.N.; Formal analysis, A.S. and H.N.; Funding acquisition, P.M.B.; Investigation, A.S. and H.N.; Methodology, A.S. and H.N.; Resources, P.M.B.; Software, A.S. and H.N.; Supervision, P.M.B., B.K.; Validation, A.S. and H.N.; Writing—original draft, A.S. and H.N.; Writing—review and editing, A.S., H.N., B.K., and P.M.B.

## Note

Supplement S3 can be found online at https://tinyurl.com/etidiliS3

