## Supplement S1 for "Drug-induced liver injury associated with elexacaftor/tezacaftor/ivacaftor from the FDA Adverse Event Reporting System (FAERS)"

Supplement S1: FAERS Outcome Definitions

Outcomes of FAERS reports are defined as follows according to the instructions for completing Form FDA 3500 (1):

**Death:** Includes only deaths that are suspected to be an outcome of the adverse event. Does not include patient deaths while using a medical product if there was no suspected association between the death and the use of the product. Does not include miscarriage or the abortion of a fetus because of a congenital anomaly (birth defect).

**Life-threatening:** The patient was suspected to be at substantial risk of dying at the time of the adverse event, or if the use or continued use of the device or other medical product might have resulted in the death of the patient.

**Hospitalization (initial or prolonged):** Admission to the hospital for one or more days (including release the same day), emergency room visit resulting in admission to hospital, or prolongation of hospitalization as a result of the adverse event. Does not include cases in which the patient in the hospital received a medical product and subsequently developed an otherwise non-serious adverse event, unless the adverse event prolonged the hospital stay. Emergency room visits that do not result in admission to the hospital should be evaluated for one of the other serious outcomes.

**Other Serious or Important Medical Events:** Event does not fit the other outcomes, but the event could have jeopardized the patient and/or required medical or surgical intervention to prevent one of the other outcomes. Examples include allergic bronchospasm requiring treatment in an emergency room, serious blood dyscrasias (blood disorders) or seizures / convulsions that do not result in hospitalization. Development of drug dependence or drug abuse would also be reported in this category.

**Required Intervention to Prevent Permanent Impairment or Damage:** It was believed that medical or surgical intervention was necessary to preclude permanent impairment of a body function, or prevent permanent damage to a body structure, either situation suspected to be due to the use of a medical product.

**Disability or Permanent Damage:** Adverse event resulted in a substantial disruption of a person's ability to conduct normal life functions. Examples include significant, persistent or permanent change, impairment, damage or disruption in the patient's body function/structure, physical activities and/or quality of life.

**Congenital Anomaly/Birth Defects:** It is suspected that patient exposure to a medical product prior to conception or during pregnancy may have resulted in an adverse outcome in the child.
