## Supplement S2 for "Drug-induced liver injury associated with elexacaftor/tezacaftor/ivacaftor from the FDA Adverse Event Reporting System (FAERS)"

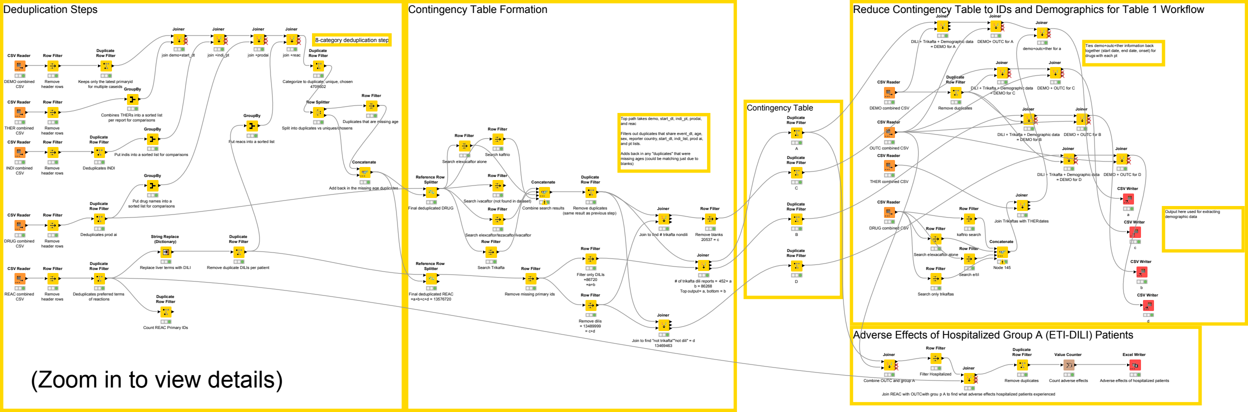

Deduplication Steps
Contingency Table Formation
Reduce Contingency Table to IDs and Demographics for Table 1 Workflow
Contingency Table
Adverse Effects of Hospitalized Group A (ETI-DILI) Patients
8-category deduplication step
CSV Reader
DEMO combined CSV
THER combined CSV
INDI combined CSV
DRUG combined CSV
REAC combined CSV
Row Filter
Remove header rows
Duplicate Row Filter
Keeps only the latest primaryid for multiple caseids
GroupBy
Combines THERs into a sorted list per report for comparisons
Deduplicates INDI
Put indis into a sorted list for comparisons
Put drug names into a sorted list for comparisons
Deduplicates prod ai
String Replace (Dictionary)
Replace liver terms with DILI
Remove duplicate DILIs per patient
Deduplicates preferred terms of reactions
Count REAC Primary IDs
Joiner
join demo+start_dt
join +indi_pt
join +prodai
join +reac
Put reacs into a sorted list
Categorize to duplicate, unique, chosen
4709602
Row Splitter
Split into duplicates vs uniques/chosens
Duplicates that are missing age
Concatenate
Add back in the missing age duplicates
Reference Row Splitter
Final deduplicated DRUG
Final deduplicated REAC =a+b+c+d = 13576720
Search elexacaftor alone
Search kaftrio
Search ivacaftor (not found in dataset)
Search elexcaftor/tezacaftor/ivacaftor
Search Trikafta
Combine search results
Remove duplicates (same result as previous step)
Top path takes demo, start_dt, indi_pt, prodai, and reac
Filters out duplicates that share event_dt, age, sex, reporter country, start_dt, indi_list, prod ai, and pt lists.
Adds back in any "duplicates" that were missing ages (could be matching just due to blanks)
Join to find # trikafta nondili
Remove blanks 20537 = c
Remove missing primary ids
Filter only DILIs =86720 =a+b
Remove dilis = 13489999 = c+d
### of trikafta dili reports = 452= a b = 86268 Top output= a, bottom = b
Join to find "not trikafta""not dili" = d 13469463
A
B
C
D
DILI + Trikafta + Demographic data = DEMO for A
DEMO+ OUTC for A
demo+outc+ther for a
Ties demo+outc+ther information back together (start date, end date, onset) for drugs with each pt
Remove duplicates
DILI + Trikafta + Demographic data = DEMO for C
DEMO + OUTC for C
DILI + Trikafta + Demographic data = DEMO for B
DEMO + OUTC for B
OUTC combined CSV
DILI + Trikafta + Demographic data = DEMO for D
DEMO + OUTC for D
Join Trikaftas with THERdates
kaftrio search
Search elexacaftor alone
Search e/t/i
Search only trikaftas
Node 145
CSV Writer
a
b
c
d
Output here used for extracting demographic data
Combine OUTC and group A
Filter Hospitalized
Join REAC with OUTCwith grou p A to find what adverse effects hospitalized patients experienced
Value Counter
Count adverse effects
Excel Writer
Adverse effects of hospitalized patients
(Zoom in to view details)

### ETI-DILI (Group A) Patient Demographics Workflow

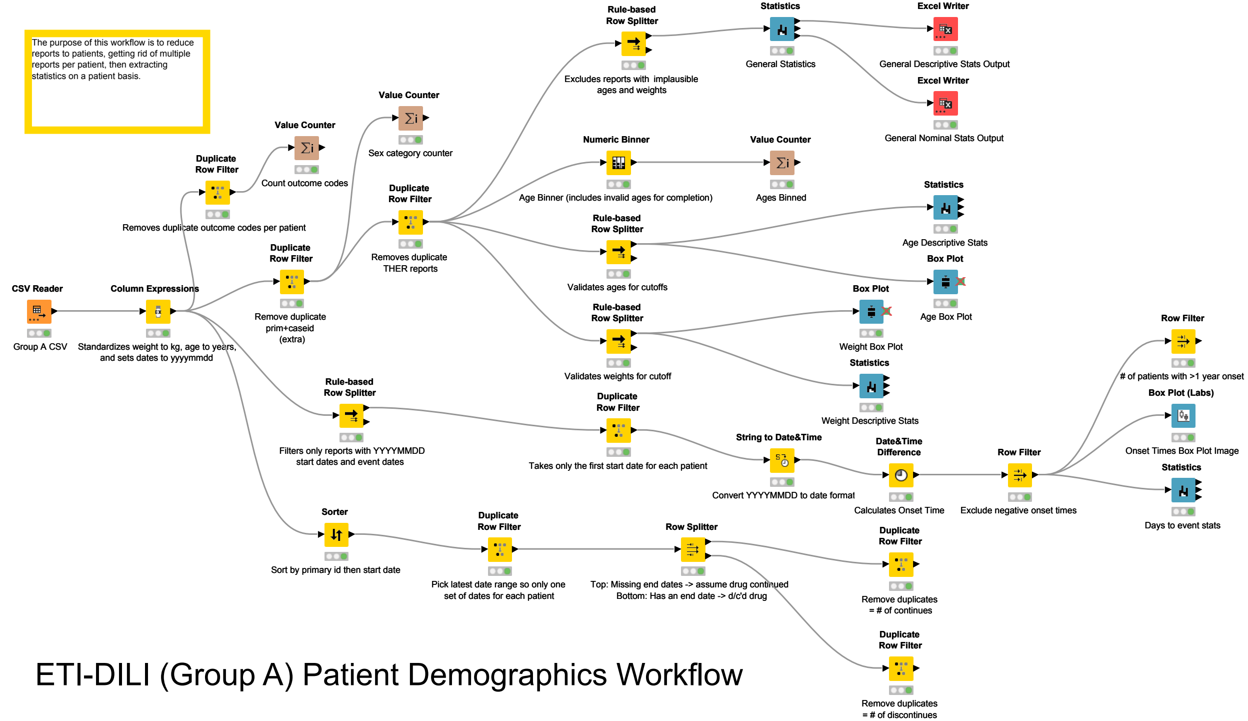

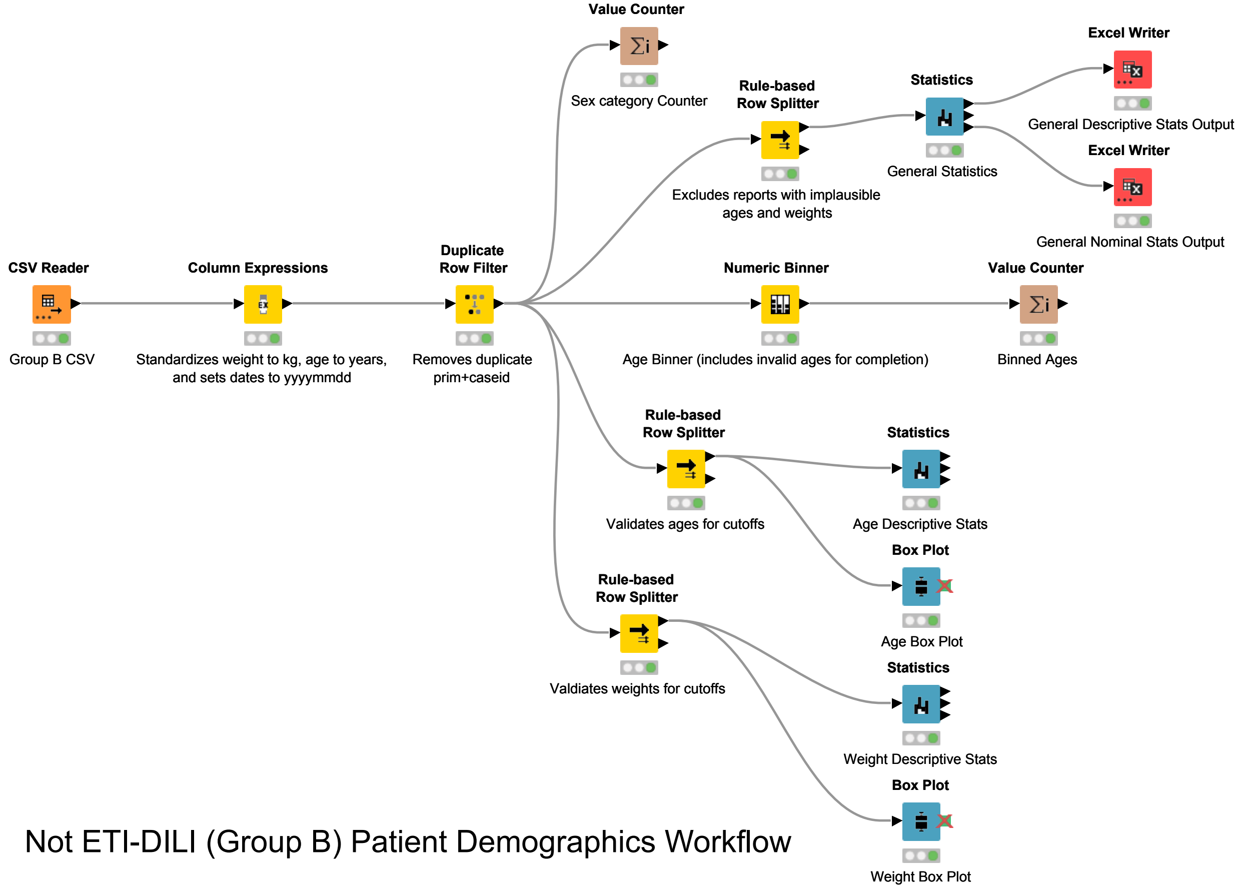


### Not ETI-DILI (Group B) Patient Demographics Workflow

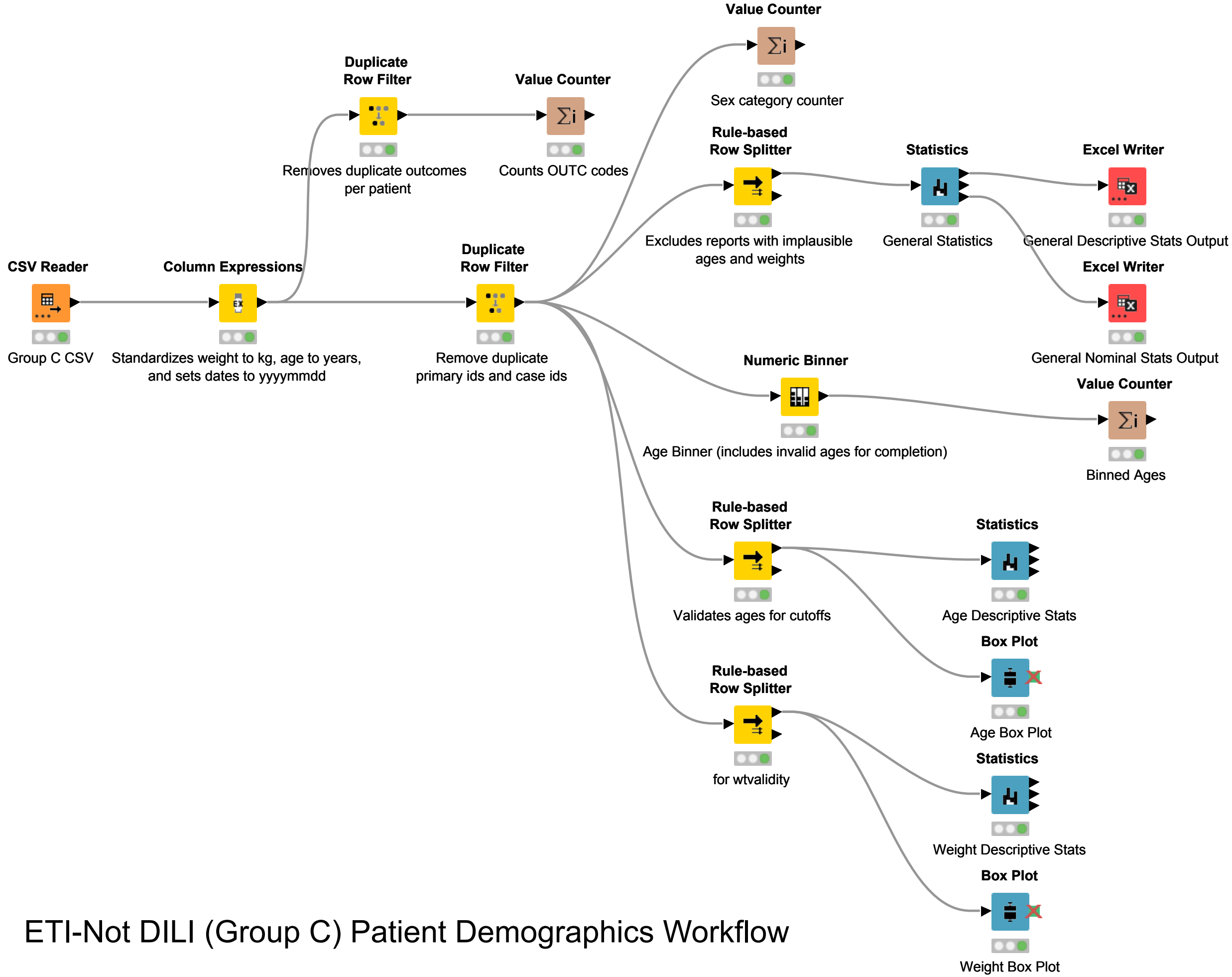


ETI-Not DILI (Group C) Patient Demographics Workflow

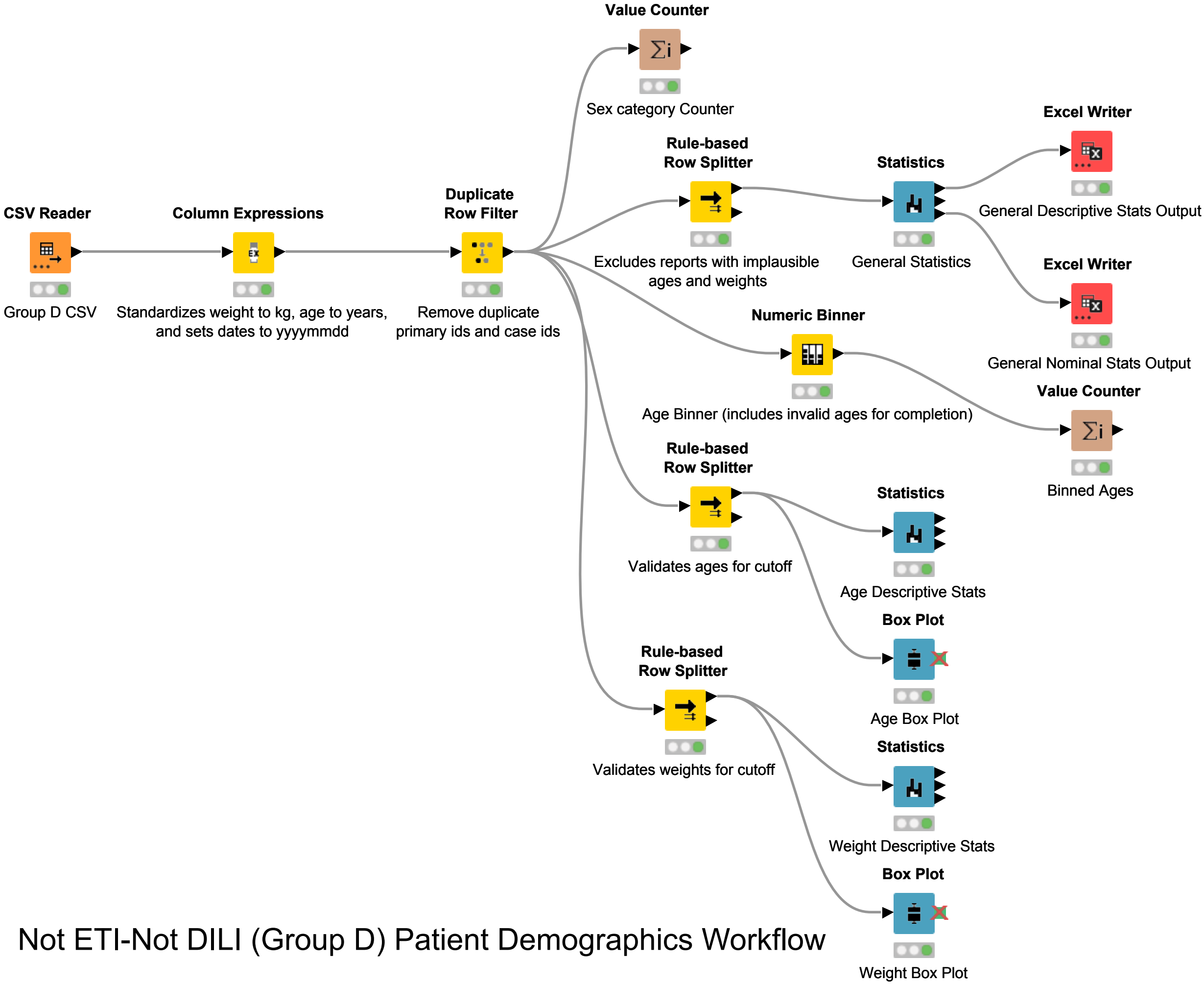


Not ETI-Not DILI (Group D) Patient Demographics Workflow
